# High prevalence of loss of Y chromosome in the spermatozoa of young cancer survivors

**DOI:** 10.64898/2026.03.20.26348822

**Authors:** Jonatan Axelsson, Bozena Bruhn-Olszewska, Daniil Sarkysian, Ellen Markljung, Monika Horbacz, Indira Pla, Aniel Sanchez, Hannah Nenonen, Angel Elenkov, Jan P. Dumanski, Aleksander Giwercman

## Abstract

Cancer-related genomic instability (GI) may cause genetic alterations in spermatozoa, implying health issues not only in cancer survivors, but also in their children [1, 2]. We therefore studied Loss of Y chromosome (LOY), considered as hallmark of GI [3-15], in spermatozoa and blood from survivors of childhood and testicular cancer (CC, TC), and controls (CTRL). We found that LOY was statistically significantly more frequent in spermatozoa from cancer survivors than in controls (Odds Ratio [OR]=2.2 for CC vs. CTRL and OR=2.4 for TC vs. CTRL). Furthermore, LOY was about an order of magnitude more prevalent in spermatozoa than in blood among 18–53-year-old males within all cohorts. Our findings suggest that LOY in spermatozoa might be a clinically useful marker of GI, reduced fertility and disease predisposition in males. Introducing LOY in spermatozoa as a biomarker opens a new research avenue into disease prevention and the causes and consequences of LOY.

---

Genomic instability (GI), being a persistent tendency of a cell or cell-lineage to acquire genetic alterations at an elevated rate compared with normal cells, is considered an important pathogenetic mechanism leading to cancer development [16]. A clearly under-researched area is whether GI in male cancer patients leads to genetic aberrations in spermatozoa, which can be transmitted to and cause health issues in their offspring. In line with the above, we have reported an increased risk of congenital malformations in children of male cancer patients, regardless of whether the conception took place before or after the cancer diagnosis and treatment [1, 2]. Furthermore, increased risk of malignant diseases was reported among children of males diagnosed with cancer [17]. We thus hypothesized that global GI might be a pathogenetic factor linking these conditions. Since LOY is a well-known marker of GI [3-15], and is rarely detectable in haematopoietic cells of men below the age of 50 [7, 18], we also aimed to explore if LOY in spermatozoa (SPERM-LOY) occurs in younger men and whether it might be a less age**-**dependent phenomenon.

LOY is a very common aneuploidy in normal human cells as well as in cancer cells and has predominantly been studied in the haematopoietic lineage (H-LOY) [13], with only a few exceptions [19-21]. H-LOY is now recognized as the most common post-zygotic aberration in males, affecting more than 40% of those over 70 years old [7, 13]. Moreover, H-LOY is associated with, or induced by, exposure to common mutagens, such as tobacco smoking, air pollution, glyphosate and arsenic, suggesting that it might function as a sensor for accumulation of various mutations leading to GI [13]. GWAS studies identified genetic predisposition for development of H-LOY and the affected genes were related to, for instance, the cell-cycle progression, the mitotic spindle-assembly checkpoint, and the DNA damage response, further emphasizing the role of LOY in GI [7]. Furthermore, H-LOY has been associated with numerous preferentially chronic diseases occurring late in life [13] and has also been shown to cause cardiac fibrosis [22] as well as driving development of cancer [15, 23]. Considering the above-described background, we assessed LOY in paired blood and semen samples in three cohorts collected from survivors of childhood cancer (CC) and testicular cancer (TC), as well as age-matched healthy and fertile controls (CTRL). Finally, we tested the hypothesis of biological link between LOY and GI, investigating prevalence of structural genomic aberrations on autosomes in spermatozoa of men with and without SPERM-LOY.

## Results

All TC patients had the diagnosis of testicular germ cell tumour, while the CC cohort had a mixture of 24 diagnoses (Supplementary Table 1). The majority of subjects (292 out of 315) provided paired semen and blood samples (Supplementary Table 2). CC and CTRL delivered one semen and one blood sample each at the same timepoint, whereas TC patients provided one blood sample and up to six semen samples collected longitudinally. Clinical data included e.g., age at diagnosis, age at sampling, smoking, clinical treatments (chemotherapy and radiotherapy), DNA fragmentation index and body mass index (BMI); these covariates and their interactions were used in statistical analyses (see below). Concentration of spermatozoa was available for all cohorts and we excluded nine TC patients with zero sperm counts, to assure that DNA from germ cells was analysed. The human semen is known to contain immune cells [24-27] and if such cells have LOY, this could introduce false-positives, especially in the absence of spermatozoa. However, our study design, with blood and semen from the same subjects, should be largely protective against this bias, because we can control if leukocytes have detectable LOY.

After DNA extraction, samples were genotyped on an Illumina SNP-genotyping array and analysed with the well-established method (Mosaic Chromosomal Alterations, MoChA) to assess LOY, as described [7, 21]. We applied strict quality control for inclusion of genotyped samples into further analyses, based on genotyping quality and visual inspection of genome-wide profiles. The final number of subjects in this study was 76, 113 and 126 for CC, TC and CTRL cohorts, respectively, giving in total 759 blood and semen samples (Table 1, Supplementary Table 2, and graphical abstract in Supplementary Fig. 1). MoChA can detect LOY starting at >1% LOY cellular fraction and this threshold was applied throughout our study. However, we also tested nine additional thresholds (from >2% to >10% LOY cellular fraction) and all these thresholds produced directionally concordant effects across cohorts (Supplementary Fig. 2 and Supplementary Fig. 3).

**Table 1.**
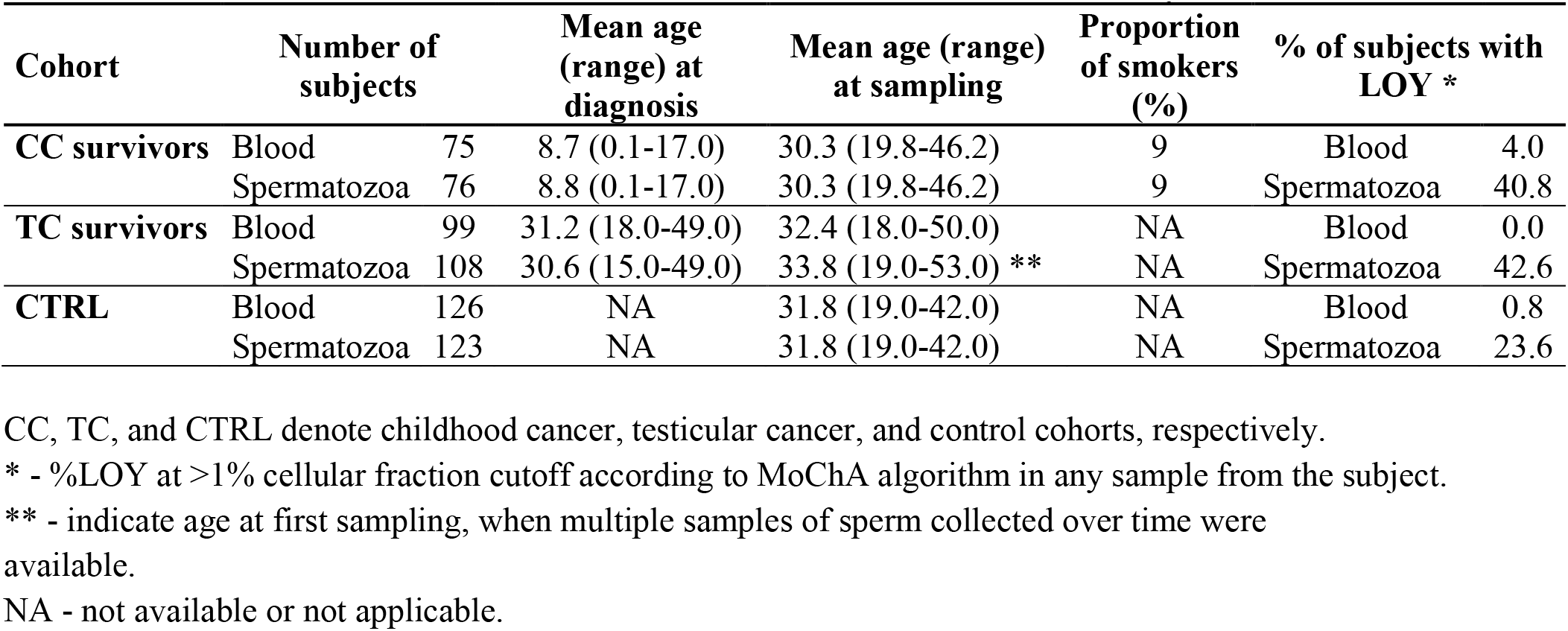
Basic characteristics of the studied cohorts and results from LOY analyses.

| **Cohort** | **Number of subjects** |  | **Mean age (range) at diagnosis** | **Mean age (range) at sampling** | **Proportion of smokers (%)** | **% of subjects with LOY \*** |  |
| --- | --- | --- | --- | --- | --- | --- | --- |
| **CC survivors** | Blood | 75 | 8.7 (0.1-17.0) | 30.3 (19.8-46.2) | 9 | Blood | 4.0 |
|  | Spermatozoa | 76 | 8.8 (0.1-17.0) | 30.3 (19.8-46.2) | 9 | Spermatozoa | 40.8 |
| **TC survivors** | Blood | 99 | 31.2 (18.0-49.0) | 32.4 (18.0-50.0) | NA | Blood | 0.0 |
|  | Spermatozoa | 108 | 30.6 (15.0-49.0) | 33.8 (19.0-53.0) ** | NA | Spermatozoa | 42.6 |
| **CTRL** | Blood | 126 | NA | 31.8 (19.0-42.0) | NA | Blood | 0.8 |
|  | Spermatozoa | 123 | NA | 31.8 (19.0-42.0) | NA | Spermatozoa | 23.6 |

CC, TC, and CTRL denote childhood cancer, testicular cancer, and control cohorts, respectively.
\* - %LOY at >1% cellular fraction cutoff according to MoChA algorithm in any sample from the subject.
\*\* - indicate age at first sampling, when multiple samples of sperm collected over time were available.
NA - not available or not applicable.

The occurrence of LOY in spermatozoa (SPERM-LOY) using >1% LOY threshold was 24% in CTRL and 43% and 41% in TC and CC groups, respectively (Table 1). The prevalence of SPERM-LOY was significantly increased in both cancer cohorts, CC vs. CTRL (odds ratio [OR]=2.2, 95% highest density interval [95%-HDI]=1.02-3.88) and TC vs. CTRL (OR=2.4, 95%-HDI=1.23-4.04) (Fig. 1). We also calculated the OR for the prevalence of SPERM-LOY in cancer survivors using combined data for CC and TC cohorts versus CTRL cohort. The OR was 2.25 with 95%-HDI=1.23-3.50 and adjusted p-value of 0.0014.

**Fig. 1.**
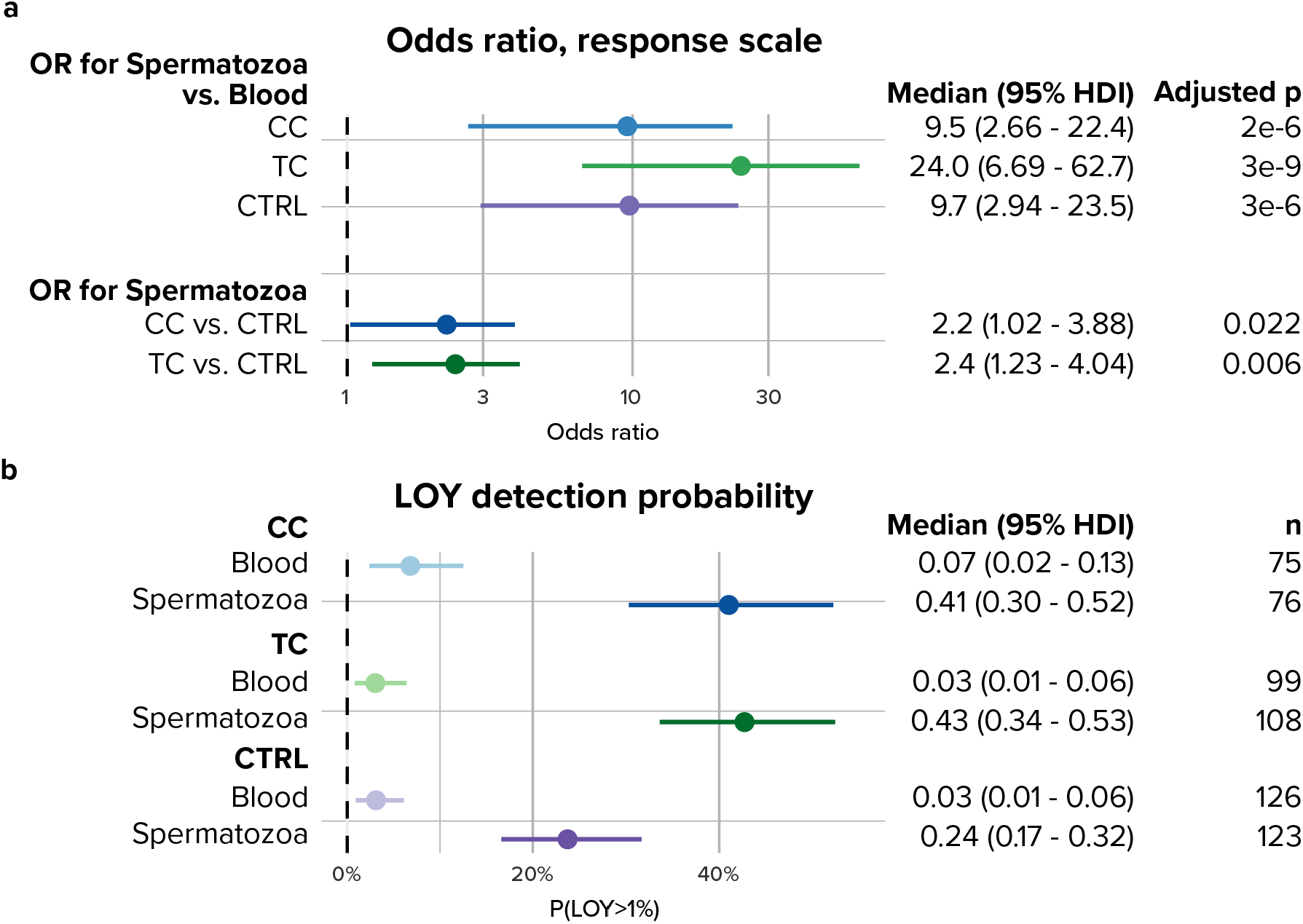
Probability and odds of detecting LOY in blood and spermatozoa in three cohorts. **a**, Corresponding odds ratio contrasts on the response scale: spermatozoa vs. blood within each cohort, as well as CC vs. CTRL and TC vs. CTRL in spermatozoa. **b**, Probabilities of detecting LOY in blood and spermatozoa of CC and TC cancer survivors and CTRLs. Points are posterior medians, horizontal bars are 95% highest-density intervals (HDI), and p-values are adjusted for multiple testing (see Statistics).

To assess whether LOY prevalence was associated with clinical parameters, such as age, cancer treatment (chemo- or radiotherapy) and pre- vs. post-pubertal cancer diagnosis and treatment, we compared the baseline and covariate-augmented models. None of them showed significant model performance improvement or effect across the cohorts (Methods and Supplementary Table 3). Moreover, for H-LOY, no association with cancer diagnosis or treatment was observed. This contrasts with the study showing that local radiotherapy resulted in a higher %LOY in saliva and blood of irradiated prostate cancer patients than in those who were not irradiated [28]. However, the median age of patients in our study was considerably lower than in the aforementioned cohort (30 vs. 72 years), which might explain this discrepancy.

Numerous studies of H-LOY in males older than 50 years showed that age has a very strong, exponential effect on the prevalence and cellular frequency of this aberration [3-5, 7, 9, 15, 18, 29, 30]. However, for males aged 18-53 years studied here, adding age as a covariate did not improve model performance and showed no meaningful effect on prevalence of LOY in blood or spermatozoa. Figure 1 shows the probabilities and odds ratios without correction for age, while Supplementary Fig. 5 illustrate corresponding analysis adjusted for age at sampling, with very similar results.

Another important finding shown in Fig. 2, and Supplementary Fig. 4 illustrates the results of LOY analyses in the three cohorts using blood vs. spermatozoa from subjects with paired samples only and also including subjects with unpaired samples (either blood or spermatozoa), respectively. As shown in these figures and the summary in Table 1, LOY was considerably more common in semen than in blood, by about an order of magnitude in the CC and CTRL cohorts, and even higher in the TC cohort. The odds ratios for these comparisons between SPERM-LOY and H-LOY are as follows: CC (OR=9.5, 95%-HDI=2.66-22.4); TC (OR=24.0, 95%-HDI=6.69-62.7); and CTRL (OR=9.7, 95%-HDI=2.94-23.5) (Fig. 1).

**Fig. 2.**
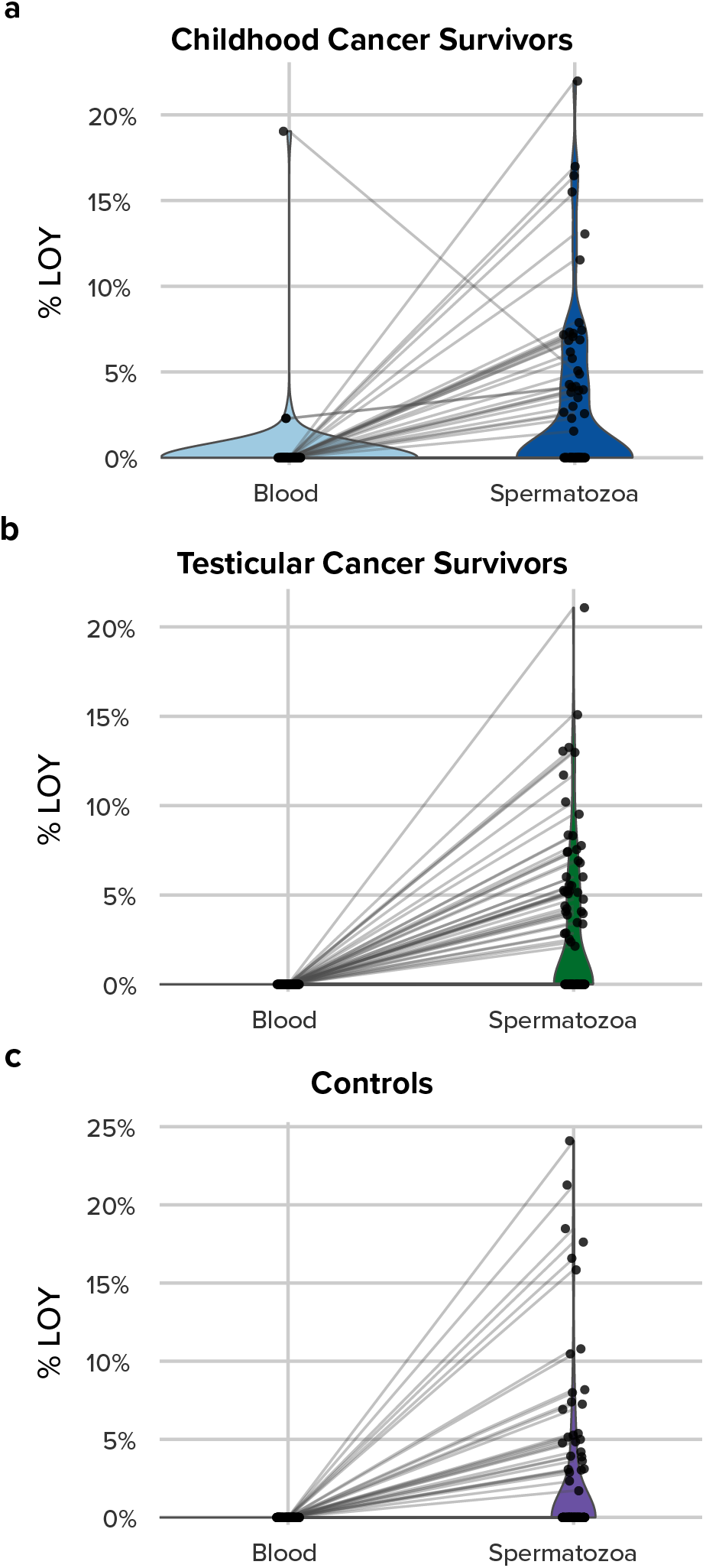
Paired comparison of LOY between blood and spermatozoa from the same subjects across three cohorts. Violin plots show the distribution of percent LOY cellular fraction (%). **a**, CC blood vs. CC spermatozoa from 75 subjects. **b**, TC blood vs. TC spermatozoa from 94 subjects. **c**, CTRL blood vs. CTRL spermatozoa from 123 subjects. Only subjects contributing both blood and one semen sample are shown here, with paired sample connected by a line to emphasize intra-individual changes. Since age had no detectable effect on LOY, for TC patients with multiple semen samples, the sample with the highest LOY was used.

Moreover, we investigated if SPERM-LOY levels were varying intra-individually. In the TC cohort, semen samples were collected up to 6 times within 60 months after orchidectomy (Supplementary Fig. 6), which allowed for a longitudinal analysis of SPERM-LOY levels within the same patient. The results are summarized in Fig. 3 and Supplementary Fig. 7, using age of the patients and time since orchidectomy on the x-axes, respectively. In conclusion, among the TC patients with two or more semen samples, SPERM-LOY varied intra-individually across the different time points with 47% of TC patients showing variation. This suggests a highly dynamic pattern of clonal expansions in spermatozoa with LOY and this number is probably an underestimate, considering that 35% of TC patients were studied using only one semen sample. Intra-individual changes in H-LOY levels have been reported before, with measurement intervals either spanning many years [3, 9, 31] or limited to 1–3 months, the latter during COVID-19 infection [32]. Thus, spermatozoa also display this feature and with a high level of variation.

**Fig. 3.**
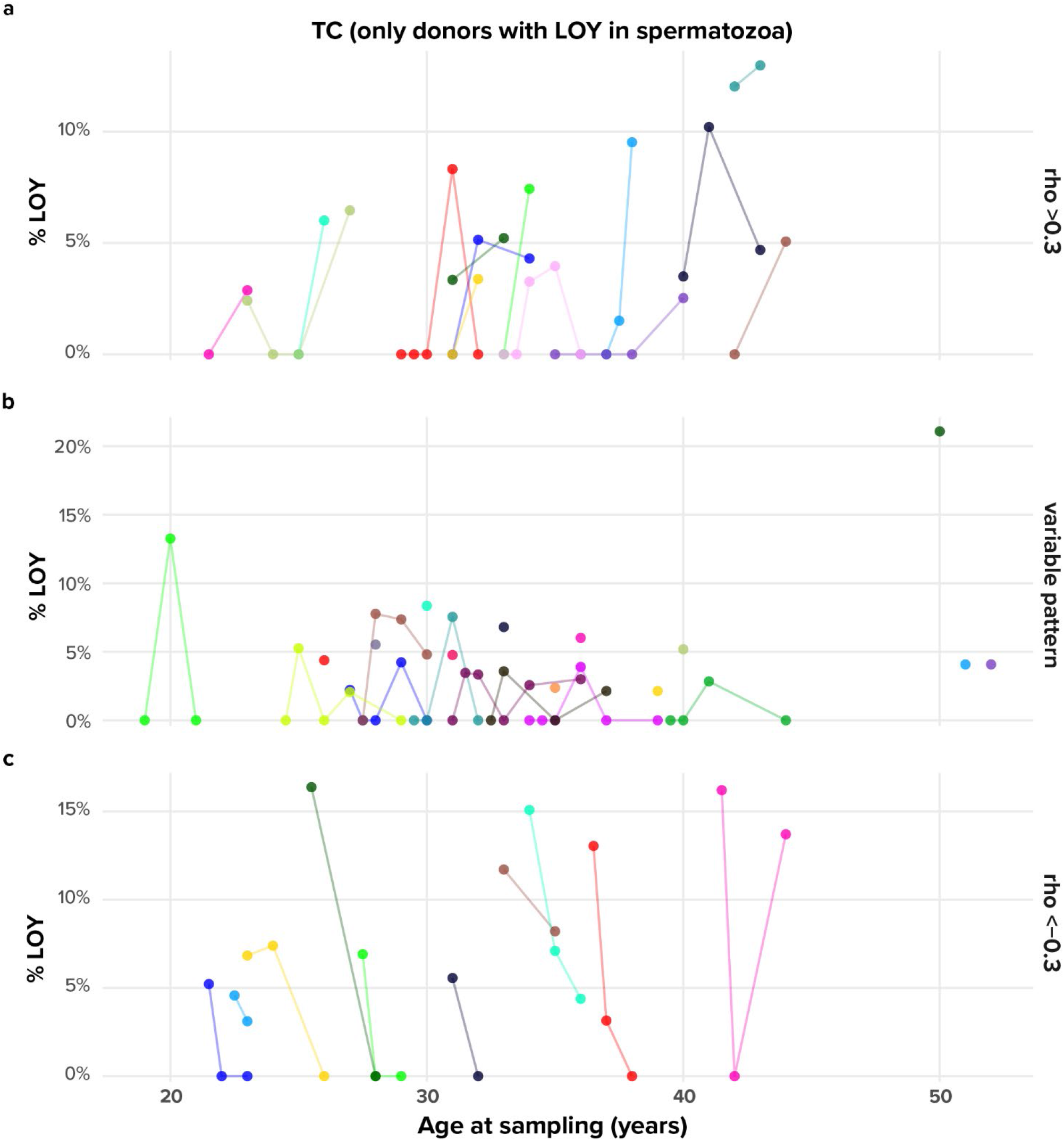
Longitudinal LOY in spermatozoa from testicular cancer (TC) survivors, plotted by age at sampling. Spaghetti plots show LOY cellular fraction (%) in spermatozoa for TC patients with at least one measurement >1% LOY. Each line is one patient. Colours distinguish patients only within a panel. Panels separate subjects by empirical trend defined from the Spearman correlation ρ between LOY and age at sampling. **a**, Increasing LOY (ρ>0.3) in 14 patients. **b**, 12 patients with varying levels of LOY across at least two samples and 10 patients with LOY, but without serial samples. **c**, Decreasing LOY (ρ<−0.3) in 10 patients.

We further tested a hypothesis whether SPERM-LOY could be associated with broader GI involving structural genomic aberrations on autosomes in germ cells. MoChA output suggested that there might be as many as 4905 autosomal chromosomal alterations (ACAs) in the 759 samples of blood and spermatozoa in 315 studied subjects. Our goal was to identify sperm-specific ACAs (SS-ACAs), defined as alterations observed in spermatozoa but absent in the paired blood sample. We used for this purpose in-house manual curation pipeline described previously [21], adapted for credible validation of SS-ACAs. In total, 506 subject-level Log R ratio/B-allele frequency/phased BAF (LRR/BAF/pBAF) profiles for blood and spermatozoa were examined manually by visual inspection, yielding 207 curated subject-level SS-ACAs, comprising 107 losses, 61 gains, 18 copy-neutral loss-of-heterozygosity (CN-LOH) calls and 21 events of undetermined type (Fig. 4). SS-ACA burden was calculated in two ways; as the number of SS-ACAs per subject and as a total SS-ACA genomic length per subject. The maximum burden was nine events in a single subject and 207 individuals did not show any. Across all subjects, this burden was significantly higher in the subjects with LOY≥1% than in the LOY<1% group (p=2E-11) (Fig. 4) and the same pattern was observed in cancer survivors and controls studied separately (Table 2). Eight representative subjects with SS-ACAs are shown in Supplementary Fig. 8. Furthermore, Supplementary Fig. 9 shows the genomic distribution of different types of curated SS-ACAs across the autosomes, which might suggest a non-random pattern. However, the cohort is too small to reliably investigate possible hotspots.

**Fig. 4.**
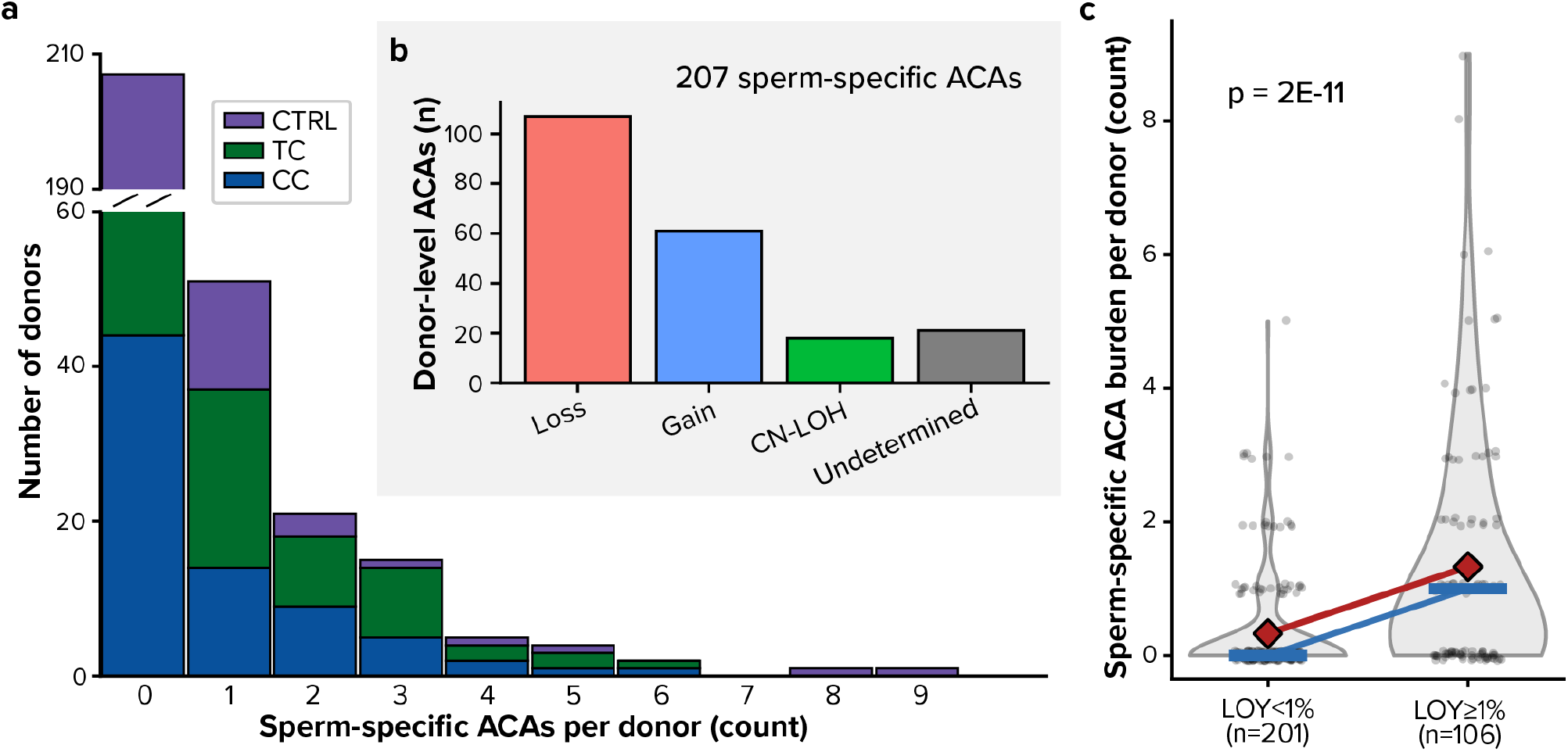
Subject-level sperm-specific autosomal chromosomal alterations (ACAs) are enriched in SPERM-LOY subjects. **a**, Distribution of subject-level sperm-specific ACAs (SS-ACAs) counts per subject, shown as a histogram stacked by cohort (CC, TC, CTRL) with y-axis break for the large zero-burden bin. **b**, Composition of 207 curated subject-level SS-ACAs by call type. **c**, Subject-level SS-ACA counts stratified by SPERM-LOY status (LOY<1% vs LOY≥1%). Violin plots show the distribution across all available subjects (p=2E-11, two-sided Mann-Whitney U test, see Methods); points represent SS-ACA counts of individual subjects (jittered for visibility). Red diamonds indicate group means, blue horizontal lines indicate medians, and the connecting lines highlight the shifts in means and medians between groups. Overlapping ACA calls from repeated sperm follow-up samples of the same subject were collapsed so that the same underlying alteration was counted once per subject (see Methods).

**Table 2.**
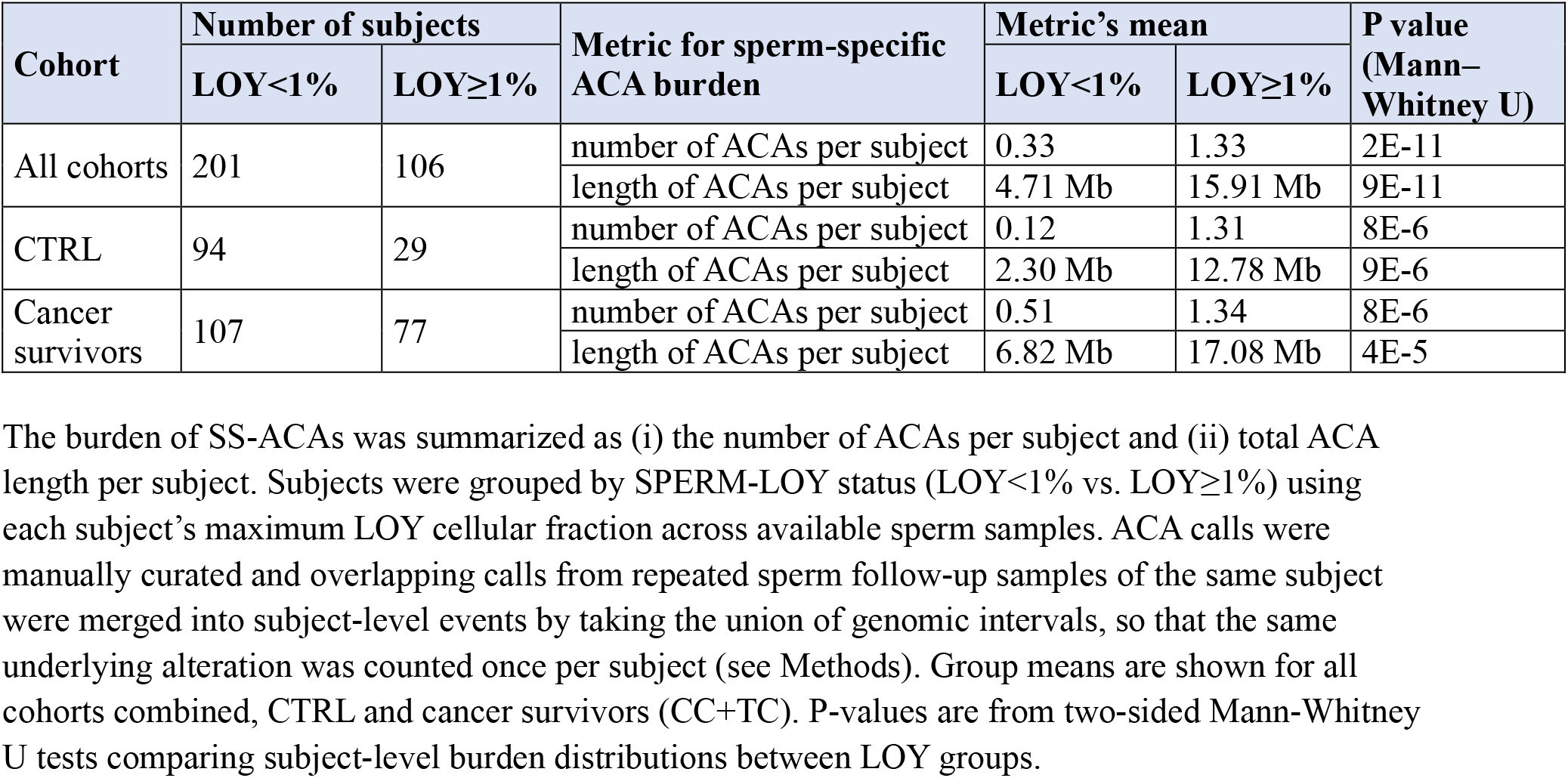
Sperm-specific autosomal chromosomal alterations (SS-ACAs) burden is higher in subjects with SPERM-LOY.

We also evaluated whether semen quality variables, such as sperm concentration and DNA Fragmentation Index (DFI), were associated with SPERM-LOY status. DFI data were unavailable for controls. In Mann–Whitney comparisons between LOY<1% and LOY≥1% groups, neither sperm concentration nor DFI differed within CC and TC cohorts or in combined analyses, and this remained unchanged across alternative SPERM-LOY thresholds from 1% to 5% (details not shown). These results make it unlikely that variation in sperm concentration or DFI explains the observed association between SPERM-LOY and SS-ACA burden.

## Discussion

Our main finding was a statistically significant increase in SPERM-LOY among the cancer survivors relative to controls. This increase was independent of the cancer treatment or whether the disease was diagnosed and treated before or above the age of 10-14 years. Before puberty, no active meiosis occurs in the seminiferous epithelium and it has therefore been hypothesized that the germ cells before puberty might be more resistant to the negative effects of cancer and its treatment on the genome [33, 34]. However, our results did not reveal any difference between pre- and post-pubertal cases, providing no support for this hypothesis. Overall, our SPERM-LOY results are in line with previous reports showing that high cell proliferation and renewal in some tissues, such as haematopoietic cells or microglia, is connected with risk to develop LOY [13]. Throughout our analyses, we assume that SPERM-LOY is due to production of haploid spermatozoa that lost their Y chromosome during meiosis, meaning that semen contains a mixture of 23X, 23Y and 22(LOY) cells. However, another, but perhaps less likely explanation is a strong imbalance between X- and Y-containing spermatozoa, with predominance of the former. This limitation of the current analysis warrants further investigation. Another open question is at which phase of meiosis SPERM-LOY occurs.

SPERM-LOY might be an expression of global GI in a subject. Assuming that this hypothesis is correct, GI in the germline could lead to production of spermatozoa containing other (than LOY) types of mutations. Our results from analyses of ACAs strongly support this notion (Fig. 4 and Table 2). This might be reminiscent of co-existence of clonal hematopoiesis (CH) mutations and H-LOY in leukocytes [9, 35]. The extension of this combined SPERM-LOY and ACAs analysis might lead to delineation of ACA hotspots, which can be transmitted to and cause health issues in the offspring. Notably, Neville *et al*. [36] has recently detected 40 genes under positive selection in the male germline that had activating or loss-of-function mechanisms and were involved in diverse cellular pathways. Many of these were associated with developmental or cancer predisposition in children. Provided that increased SPERM-LOY is associated with GI involving ACAs in spermatozoa, this could explain the increased frequency of congenital malformations in children of fathers that survived cancer [1], and also being found in children conceived before the paternal cancer diagnosis [2]. The CTRL cohort showed SPERM-LOY in ∼20% of cases and the intriguing question is whether the children of these subjects are at higher risk of congenital malformations and whether these men themselves are predisposed to cancer or other age-related diseases later in life? On the other hand, a certain transient, low threshold level of SPERM-LOY might be phenotypically neutral and represent normal post-zygotic genetic variation and future studies should address this issue.

The major focus of previous LOY studies has been on haematopoietic cells; using whole blood, sorted leukocytes, or in single-cells [3, 5, 13, 32, 37]. LOY has been reported in several other normal tissues; for instance, buccal mucosa, atherosclerotic plaques and normal urothelium, albeit at a lower frequency [19-21]. The buccal mucosa (essentially free from leukocytes) had considerably less LOY than blood from the same subjects [20]. Our results suggest that a high-level of SPERM-LOY might be, from a clinical point of view, a more feasible and better method (than H-LOY) for identifying subjects with increased risk of GI-related diseases. We should mention here a recent forensic genetic study that measured SPERM-LOY in 12 young males (along with a higher number of blood and saliva samples) and state that semen may not be a proper substrate to study LOY in the context of age estimation [38]. We believe that, considering the small number of studied subjects, this conclusion might be premature.

Variation in SPERM-LOY levels across sampling timepoints may reflect cancer-related GI, which could be manifested as a transient DNA damage in progenitors leading to spermatozoa [39]. However, LOY might also be causally involved in the development of TC by promoting a proliferation of cancer progenitor cells in the testes resulting in a testicular germ cell tumour, as it has already been shown for several other cancer forms [15, 23].

Our CC cohort shows a considerable heterogeneity of cancer diagnoses (Supplementary Table 1). Thus, future studies should test the strength of associations between SPERM-LOY and disease development in a larger cohort of cancer survivors with less diversified diagnoses. Notably, incident childhood cancer cases are 28% higher in boys than in girls [40] and this is consistent with a strong cancer bias for older males [13, 41-43]. The majority of tumour diseases in children fall into two categories: *i)* haematopoietic cancer such as acute lymphocytic leukaemia and other less common forms of haematopoietic malignancies (acute myeloid leukaemia; non-Hodgkin and Hodgkin lymphomas); and *ii)* brain and peripheral nervous system tumours^32^. Moreover, as previously shown, H-LOY has been associated with about 200 SNPs that predispose to this aneuploidy^3-5^. Thus, our results also raise the possibility of a genetic predisposition to SPERM-LOY, as it remains unknown whether a similar or distinct set of SNPs might predispose to SPERM-LOY. This question could be answered assuming the availability of large cohorts of men who provide semen and blood for such studies. We are tempted to speculate that SPERM-LOY might be a precursor of H-LOY that will develop later in life and that might lead to diseases in males.

LOY has only been described as a mosaic event [13] and it apparently cannot exist in an inherited form affecting all somatic cells, as illustrated by Turner syndrome (45,X). Accordingly, a human foetus composed only of 45,X cells is not viable and live-born 45,X females always have a rescue cell line, predominantly with a normal karyotype [44]. Thus, if a male with a high frequency of SPERM-LOY is conceiving a new pregnancy, a 45,X foetus will not develop to term, and this might reduce his fertility in general as well as result in a lower frequency of male offspring. Of note, more girls are born to fathers surviving testicular cancer [45, 46]. This is in contrast to the general population of human births, which are male-biased, with males making up around 51–52% of live births, meaning about 105–107 male births per 100 female births. This pattern has been remarkably consistent across different populations and time periods wherever reliable birth records exist [47-49].

In conclusion, SPERM-LOY is considerably more common among young cancer survivors than in controls. Furthermore, LOY in male gametes is significantly more frequent than it is in leukocytes of young adult males. Our results suggest that LOY might be involved in the pathogenesis of childhood cancer, testicular cancer and other diseases later in life, warranting further studies. SPERM-LOY may also affect male fertility and may be the reason for the predominance of girl offspring from survivors of testicular cancer [45, 46]. Our results open a new research avenue into the causes and consequences of LOY in younger males, that are understudied with regard to this aneuploidy.

## Methods

### Study cohorts and extraction of DNA

We collected blood and semen samples from survivors of childhood cancer (CC) and testicular cancer (TC) as well as age-matched healthy and fertile controls (CTRL), in total 315 subjects. Relevant clinical data were also collected, such as age at diagnosis, age at sampling, smoking, clinical treatments (chemotherapy and radiotherapy), BMI and counts of spermatozoa. The collection was performed at Reproductive Medicine Centre, Skåne University Hospital, Malmö, Sweden. The study was approved by the local ethical committee and all participants provided written informed consent for participation in this study. DNA extraction was performed using commercially available kits. Spermatozoa and blood DNA were extracted with the genomic DNA isolation kit from Norgen (Thorold, Canada) or with PureLink™ Genomic DNA Mini Kit from ThermoFisher (Waltham, Massachusetts, USA), respectively. DNA was diluted in water or TE buffer (10 mM Tris HCl, pH 8, 0.1 mM EDTA) and stored at –80 °C.

### Genotyping and detection of LOY by MoChA from SNP array data

SNP&SEQ Technology Platform, Uppsala, Sweden, part of the National Genomics Infrastructure (NGI) Sweden and Science for Life Laboratory performed genotyping for the CC, TC and CTRL cohorts using Infinium Global Screening Array Multiethnic Disease Version 3.0 (GSAMD-24v3-0-EA_20034606). The output was filtered as described previously to remove samples indicating contamination and sex discordance, applying strict quality control for inclusion of genotyped samples in downstream analyses, based on genotyping quality and visual inspection of whole genome profiles [21]. Intensity data were processed via Mosaic Chromosomal Alterations (MoChA) according to authors’ instructions [50]. MoChA pipeline was then used for detection of Loss of Chromosome Y (LOY) as described [7, 21].

### Statistics

Statistical analysis was performed in *R* 4.5.0 [51]. To compare LOY prevalence between tissues (sperm vs. blood) and across CTRL, CC and TC cohorts, we fitted a Bayesian mixed-effects logistic regression using *brms* 2.22.0 [52] interface to *Stan* 2.32.7 [53]. We used weakly informative priors of normal(0, 1) for all fixed effects and student_t(3, 0, 2.5) for the random-effect standard deviation, following Stan/BRMS recommendations for logistic models with sparse positives [54]. Predictors were centered and scaled. Models had no intercepts with indexing approach to predictors [55]. Models were run with 4 chains, 12,000 iterations per chain (6,000 warmup), adapt_delta 0.99 and max_treedepth 15; all achieved 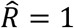 and large effective sample sizes.

For each LOY threshold *t* from 1% to 10% we created a binary indicator LOY_t_ = 1 if LOY cellular fraction exceeded *t*/100. Model families (Supplementary Table 3) compared baseline vs. covariate-augmented versions using the PSIS-LOO expected log-predictive density (ELPD) to test whether covariates or their interactions improved predictive performance. Posterior marginal probabilities for each tissue–cohort combination were obtained with *emmeans* 1.11.1 [56] using type = “response” and re_formula = NA. Spermatozoa vs. blood and cancer survivors vs. healthy controls contrasts were estimated with contrast function from *emmeans* using method = “trt.vs.ctrl”. We also reported asymptotic p-values from *emmeans* with multivariate-t adjustment within appropriate test families; these appear together with the Bayesian medians and 95% highest density intervals in the figure. Sensitivity analyses across thresholds 1–10% used the same formula, priors and settings.

### Curation of autosomal MoChA calls and subject-level sperm-specific ACA burden estimation

Autosomal MoChA calls were curated using our in-house manual curation pipeline described previously [21], adapted here to identify sperm-specific rather than post-zygotic autosomal chromosomal alterations (ACAs). In brief, intensity data were processed via MoChA WDL version 2024-05-05 according to author’s instructions and recommended quality check procedures. The output was filtered to remove samples with call rate <0.97. The decision to remove or keep each sample with BAF auto-correlation >0.03, which is indicative of sample contamination or sex discordance, was made after manual examination of hybridization quality using whole genome LRR and BAF plots. ACA calls of <10 bp were removed. The 969 of ACAs with log-odds score of ≥10 for the model based on BAF and genotype phase as well as ACAs with minimal reciprocal overlap ≥0.25 with those were manually curated by inspecting resulting 506 subject-level LRR / BAF / phased BAF profiles. We defined sperm-specific ACAs (SS-ACAs) as curated autosomal alterations observed in spermatozoa but absent in the paired blood sample or observed in some, but not all spermatozoa samples, where longitudinal samples were available. This resulted in 207 curated SS-ACAs. To avoid over-counting repeated semen follow-up samples from the same subject, ACA calls from the same subject showing minimal reciprocal overlap ≥0.25 contributed one subject-level SS-ACA event to the count burden, with event’s length defined as the length of the union of genomic intervals across such reciprocally overlapping ACAs in spermatozoa. Subject-level SS-ACA burden was summarized as: (i) the number of subject-level SS-ACAs; and (ii) the total genomic length of these SS-ACAs per subject. Subjects without SS-ACAs were retained and assigned zero burden. Group comparisons between SPERM-LOY strata were performed using two-sided Mann–Whitney U tests. P values were adjusted for multiple testing within each family of tests using the multivariate-t approach implemented in R package *emmeans*. In addition, we used two-sided Mann–Whitney U tests to compare whether sperm concentration and DNA Fragmentation Index differed between LOY<t and LOY≥t groups for t=1%, …, 5% within each cohort and in pooled strata, but none of these additional comparisons were significant after multiple testing correction.

## Data availability

Consented data that can be released are included in this article and its supplementary file. Although raw SNP genotyping data are restricted to protect patient privacy, de-identified data are available. Researchers can gain access to de-identified data by contacting the corresponding author and stating the type of data and the purpose of their data request, which will be reviewed in accordance with Research Data Governance. Researchers will be required to execute a data sharing agreement.

## Code availability

Custom code developed in this study is available at GitHub (available after the manuscript’s acceptance in a peer-reviewed journal).

## Acknowledgements

We thank Dr. Eva Tiensuu-Janson, Dr. Arja Harila Dr. Natalia Filipowicz, Dr. Jessica Nordlund and Dr. Ingrid Glimelius for critical review of the manuscript. This study was supported by funding from Swedish Cancer Society, Swedish Research Council-Medicine, Uppsala University, Medical University of Gdansk and National Science Centre – Poland to JPD; and by funds from Swedish Cancer Society, Swedish Childhood Cancer Society, Swedish Governmental Fund for Clinical Research (ALF), Interreg EU fund; MAS Cancer Fund to AG. JA was partly supported by funding from Swedish Governmental Fund for Clinical Research (ALF); young investigator award. SNP genotyping was performed by the SNP&SEQ Technology Platform in Uppsala, Sweden. The SNP&SEQ Technology Platform is part of the Science for Life Laboratory at Uppsala University and is supported as national infrastructure by the Swedish Research Council.

